# Changes in the lipidome in type 1 diabetes following low carbohydrate diet: a randomized crossover trial

**DOI:** 10.1101/2020.09.17.20196394

**Authors:** Naba Al-Sari, Signe Schmidt, Tommi Suvitaival, Min Kim, Kajetan Trošt, Ajenthen G. Ranjan, Merete B. Christensen, Anne Julie Overgaard, Flemming Pociot, Kirsten Nørgaard, Cristina Legido-Quigley

**Affiliations:** Steno Diabetes Center Copenhagen, Denmark; Danish Diabetes Academy, Denmark; Dept. of Endocrinology, Copenhagen University Hospital Hvidovre, Denmark; Dept. of Clinical Medicine, University of Copenhagen, Denmark; Institute of Pharmaceutical Science, King’s College London, UK

**Keywords:** Biomarker, Cardiovascular disease, Dyslipidaemia, Lipidomics, Low carbohydrate diet, Randomized trial, Type 1 diabetes

## Abstract

**Aims/hypothesis:** Lipid metabolism might be compromised in type 1 diabetes and the understanding of lipid physiology is critically important. This study aimed to compare the change in plasma lipid concentrations during carbohydrate dietary changes in individuals with type 1 diabetes and identify links to early-stage dyslipidaemia. We hypothesized that: (1) the lipidomic profiles after ingesting low or high carbohydrate diet for 12 weeks would be different; and (2) specific annotated lipid species could have significant associations with metabolic outcomes.

**Methods:** Ten adults with type 1 diabetes (mean±SD: age 43.6±13.8 years, diabetes duration 24.5±13.4 years, BMI 24.9±2.1 kg/m^2^, HbA_1c_ 57.6±2.6 mmol/mol) using insulin pumps participated in a randomized 2-period crossover study with a 12-week intervention period of low carbohydrate diet (< 100 g carbohydrates/day) or high carbohydrate diet (> 250 g carbohydrates/day) respectively, separated by a 12-week washout period. A large-scale non-targeted lipidomics was performed with mass spectrometry in fasting plasma samples obtained before and after each diet intervention. Longitudinal lipid levels were analysed using linear mixed-effects models.

**Results:** In total, 289 lipid species were identified from 14 major lipid classes (triacylglycerides, phosphatidylcholines, phosphatidylethanolamines, hexosyl-ceramide, sphingomyelins, lyso-phosphatidylcholines, ceramides, lactosyl-ceramide, lyso-phoshatidylethanolamine, free fatty acids, phosphatidylinositols, phosphatidylglycerols, phosphatidylserines and sulfatides). Comparing the two diets, 11 lipid species belonging to sphingomyelins, phosphatidylcholines and LPC(O-16:0) were changed. All the 11 lipid species were significantly elevated during low carbohydrate diet. Two lipid species were most differentiated between diets, namely SM(d36:1) (β±SE: 1.44±0.28, *FDR* = 0.010) and PC(P-36:4)/PC(O-36:5) (β±SE: 1.34±0.25, *FDR* **=** 0.009) species. Poly-unsaturated PC(35:4) was inversely associated with BMI and positively associated with HDL-cholesterol (*p* < 0.001).

**Conclusion/interpretation:** Lipidome-wide outcome analysis of a randomized cross-over trial of individuals with type 1 diabetes following a low carbohydrate diet showed an increase in sphingomyelins and phosphatidylcholines which are thought to reduce dyslipidaemia. The poly-unsaturated phosphatidylcholine 35:4 was inversely associated with BMI and positively associated with HDL-cholesterol (*p* < 0.001). Results from this study warrant for more investigation on the long-term effect of single lipid-species in type 1 diabetes.

**Trial registration:** Clinicaltrials.gov <u>NCT02888691</u>

**Graphical abstract:** 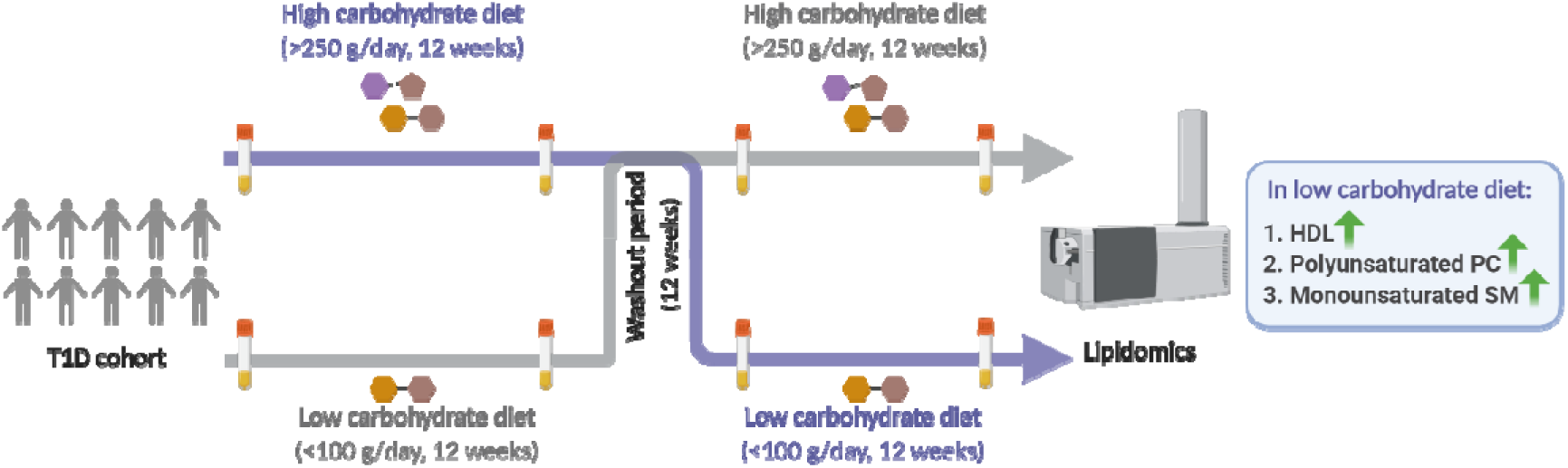

Research in context

What is already known about this subject?

- Individuals with type 1 diabetes have an increased rate of cardiovascular disease for which dyslipidaemia is a major risk factor.
- Dysregulated lipid metabolism is recognized as an established risk factor in cardiovascular diseases.

What is the key question?

- Which specific circulating lipid species are changed after 12 weeks of low- and –high carbohydrate diet and do they reflect dyslipidaemia risk?

What are the new findings?

- Plasma from individuals with type 1 diabetes showed a significant increase in phosphatidylcholine and sphingomyelin lipid species during low carbohydrate diet (n=11 lipid species).
- Poly-unsaturated phosphatidylcholine 35:4 was inversely associated with BMI and positively associated with HDL-cholesterol (*p* < 0.001).

How might this impact on clinical practice in the foreseeable future?

- This study demonstrates that unsaturated phosphatidylcholines and monounsaturated sphingomyelins were elevated with diet in individuals with type 1 diabetes following low carbohydrate diet. This points to a need for lipid-specific dietary guidelines regarding fat intake to support individuals with type 1 diabetes.

## Introduction

It is now recognized that type 1 diabetes is a major risk factor for developing cardiovascular events [1-3]. Dyslipidaemia is a modifiable risk factor for cardiovascular disease and highly prevalent in type 1 diabetes [4-6]. The present approach to diagnosing dyslipidaemia is based on the clinical measurement of the three main types of serum lipids, namely HDL-cholesterol, LDL-cholesterol and triglycerides [7]. However, since HDL-cholesterol metabolism might be compromised in type 1 diabetes, the identification of alternative biomarkers is critically important [3-6].

Lipidomics studies suggest lipids as indicators or predictors for dyslipidaemia and cardiovascular disease [8-14]. However, to date the plasma lipidome from individuals with type 1 diabetes has not been interrogated to understand diet-dependent changes in a clinical setting.

Lipidomics expands the lipid information of the three main traditional mentioned lipids, since there are many more lipid species that can be measured with modern lipidomics methods [15-17]. Lipidomics provides a profile of hundreds of individual lipid species and on low-abundance lipid species from a biological sample. Therefore, there is the potential to uncover novel insights into the pathophysiology in individuals with type 1 diabetes. The composition of the lipidome can reveal a fingerprint in relation to dyslipidaemia and cardiovascular disease in type 1 diabetes, thus it can be an important tool for early screening.

In this lipidomics analysis of a randomized cross-over trial, we compared the plasma lipids concentration from individuals with type 1 diabetes before and after ingesting isocaloric low carbohydrate diet (LCD) and high carbohydrate diet (HCD) for 12 weeks (<100g vs. >250g) separated by a 12-week washout period obtained from a previous published study [18] [<u>NCT02888691</u>]. We hypothesized that: (1) the lipidomics profiles before and after ingesting low or high carbohydrate diet for 12 weeks will be different; and (2) specific annotated lipid species have significant associations to metabolic characteristics.

## Methods

### Clinical trial

Fasting plasma samples for lipidomics analysis were collected from a previously published study [17].). Participants were adults with insulin pump-treated type 1 diabetes (mean±SD: age 43.6±13.8 years, diabetes duration 24.5±13.4 years, BMI 24.9±2.1 kg/m^2^, HbA_1c_ 57.6±2.6 mmol/mol). They underwent two 12-week diet interventions separated by a 12-week washout. Fasting plasma samples were collected before and after each intervention.

Main study outcomes from the previous article for the 14 study participants are given in **Table 1**. In brief, the study showed that glycemic variability and time spent in hypoglycemia were lower during LCD than during HCD. Mean glucose, however, was the same. Total daily bolus insulin doses were lower during LCD reflecting the lower carbohydrate intake. Finally, weight decreased during LCD, whereas it increased during HCD with a significant difference in change between groups. Systolic and diastolic BP increased during HCD (not significant), however the between-group differences were insignificant. The between-group difference in HDL-cholesterol levels were significant (p=0.005) with the LCD showing an increase (LCD: 0.06 mmol/L; HCD:-0.03 mmol/L). No changes in fasting LDL-cholesterol and triglyceride were detected [18]. In the present study, we therefore investigated whether the diet-induced changes in relevant clinical variables (BMI, BP and HDL-cholesterol) were associated with specific lipid species.

**Table 1.**
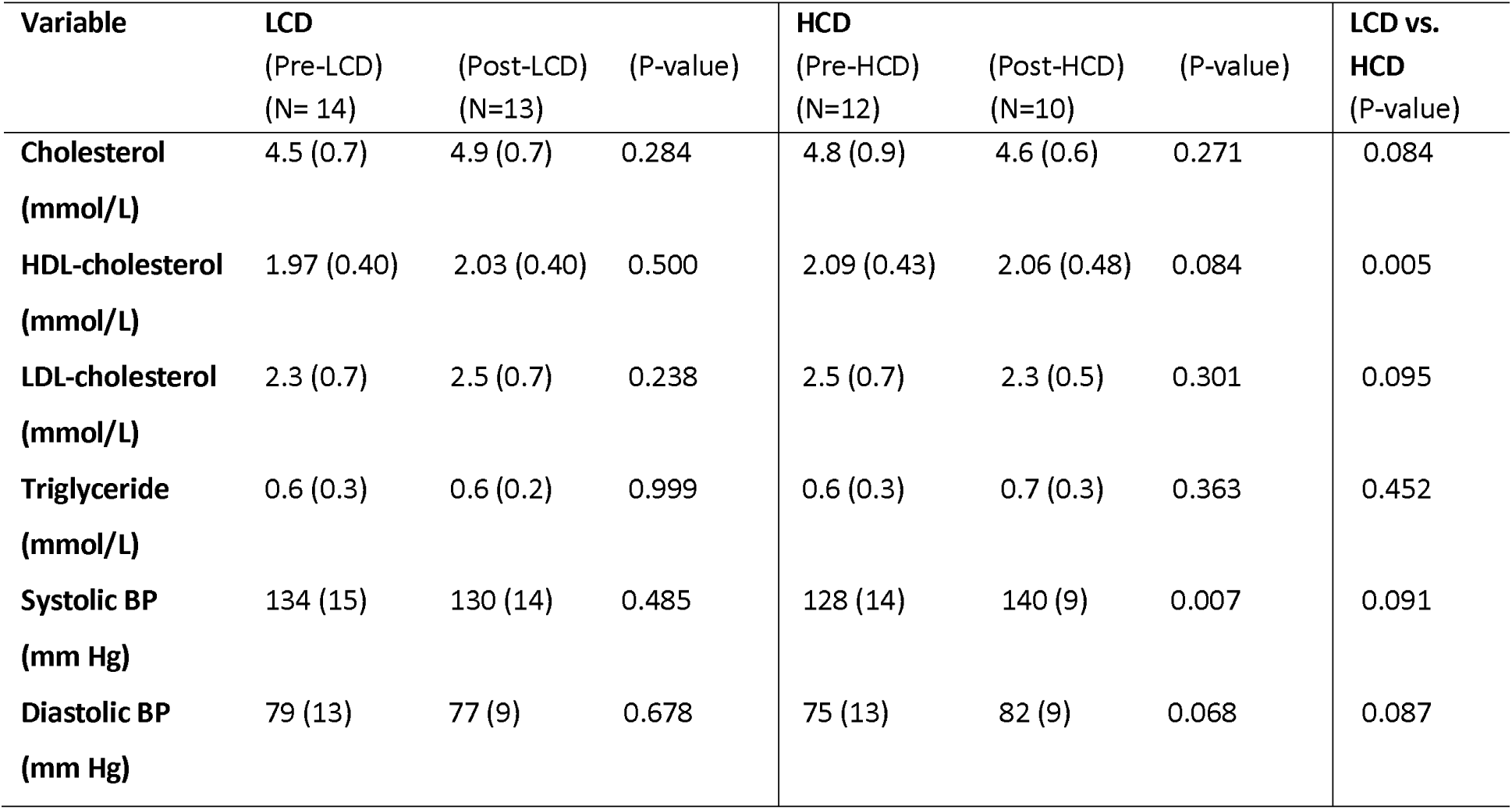
Metabolic characteristic measures before and after 12 weeks of LCD and HCD from the original study [17]. Data are mean (SD).

| Variable | LCD |  |  | HCD |  |  | LCD vs. HCD (P-value) |
| --- | --- | --- | --- | --- | --- | --- | --- |
|  | (Pre-LCD)<br>(N= 14) | (Post-LCD)<br>(N=13) | (P-value) | (Pre-HCD)<br>(N=12) | (Post-HCD)<br>(N=10) | (P-value) |  |
| **Cholesterol**<br>(mmol/L) | 4.5 (0.7) | 4.9 (0.7) | 0.284 | 4.8 (0.9) | 4.6 (0.6) | 0.271 | 0.084 |
| **HDL-cholesterol**<br>(mmol/L) | 1.97 (0.40) | 2.03 (0.40) | 0.500 | 2.09 (0.43) | 2.06 (0.48) | 0.084 | 0.005 |
| **LDL-cholesterol**<br>(mmol/L) | 2.3 (0.7) | 2.5 (0.7) | 0.238 | 2.5 (0.7) | 2.3 (0.5) | 0.301 | 0.095 |
| **Triglyceride**<br>(mmol/L) | 0.6 (0.3) | 0.6 (0.2) | 0.999 | 0.6 (0.3) | 0.7 (0.3) | 0.363 | 0.452 |
| **Systolic BP**<br>(mm Hg) | 134 (15) | 130 (14) | 0.485 | 128 (14) | 140 (9) | 0.007 | 0.091 |
| **Diastolic BP**<br>(mm Hg) | 79 (13) | 77 (9) | 0.678 | 75 (13) | 82 (9) | 0.068 | 0.087 |

All participants were given personalized advice by a dietitian focused on eating strategies for meeting the carbohydrate criteria. LCDs contained less than 100 g carbohydrates per day, and HCDs contained a minimum of 250 g carbohydrates per day. Suggestions for a healthy composition of carbohydrates, fat and protein sources were provided, however only the amount of carbohydrates was fixed. Carbohydrates intake was recorded in grams in the participants’ insulin pumps on a meal-by-meal basis, but there was no registration of actual food intake [18].

In this study, only samples from the 10 participants who completed both intervention periods are included.

### Lipidomics analysis

A modified Folch lipid extraction method was used to analyze the total lipids from plasma samples [19]. Briefly, plasma samples were randomized and lipids were extracted from 10 µL plasma using chloroform:methanol (2:1 v/v) method following addition of nine different internal standards (stable isotope labelled and non-physiological lipid species). A detailed name list of the used internal standards is available in the **Supplementary Methods 1**. Samples were analyzed in random order in positive and negative electrospray ionization modes using ultra-high-performance liquid chromatography-quadrupole time-of-flight mass spectrometry (UHPLC-Q-TOF-MS). The UHPLC system was from Agilent Technologies (Santa Clara, CA, USA) and was used as previously described [20,21].

### Data pre-processing

The lipidomics mass spectrometry data were pre-procced in MZmine 2.18.2 [22]. The workflow includes raw data import, filtering, peaks detection, chromatogram building, chromatogram deconvolution, peak list de-isotoping, peak list alignment, gap filling and finally, peak annotation was carried out by combining MS and retention time information with in-house lipid library with an m/z tolerance of 0.006 m/z and RT tolerance of 0.2 min. Lipidomics data post-processing and analysis were performed in R programming language for statistical computing (https://www.r-project.org/)[23]. The pre-processed data were normalized to internal standards. The missing values in the lipidomics dataset were imputed with the k-nearest neighbor algorithm [24]. To achieve normal-distribution, data were log-transformed. Coefficient of variation (Relative Standard Deviation; %RSD) for peak areas and retention times of lipid-class specific internal standards were calculated. The measurement of 14 lipid classes were calculated by summing the individual lipid species within each class.

### Statistical analysis

Data were analyzed and visualized in R. Lipid species-wise mixed-effect models were used to consider the four repeated measures from each participant in the cross-over trial. All statistical tests were corrected for multiple testing using the Benjamini-Hochberg method [25]. Tests were corrected for multiple comparisons with false discovery rate (FDR).

First, differences between the two diets were modelled with lipid species-wise mixed-effect models, with time, diet and time-diet interaction as fixed effects and the participant ID as random effects. We further conducted the lipid concentration changes found from the mentioned analysis within groups.

Second, associations between key lipid levels and the clinical covariates of interest were assessed: BMI, HDL-cholesterol, systolic BP and diastolic BP. In this analysis, only lipid species with significant difference between-groups from the first analyses were included. Association was tested by adding one clinical covariate at a time as an independent variable to the lipid-wise mixed effect model detailed in the first step. A detailed description about the data analysis plan is available in the **Supplementary Methods 2** Finally, lipids of interest with significant association to clinical variables was semi-quantified (relative quantified) and visualized in boxplots grouped by diets and scatterplots.

## Results

### Annotation of lipid species in plasma from type 1 diabetes

From the large-scale untargeted lipidomics analysis, lipidome-wide outcomes of the randomized trial resulted in annotation of 298 individual lipid species from 14 major lipid classes, including triacylglycerides (TGs), phosphatidylcholines (PCs), phosphatidylethanolamines (PEs), hexosyl-ceramides (HexCer), sphingomyelins (SMs), lyso-phosphatidylcholines (LPCs), ceramides (Cers), lactosyl-ceramides (LactCers), free fatty acids (FAs), phosphatidylinositols (PIs), phosphatidylglycerols (PGs), lyso-phosphatidylethanolamines (LPEs), phoshtatidylserines (PSs) and sulfatides (SHexCer). PCs and TGs dominated the data with 83 and 72 lipid species each followed by SMs and PEs with 30 identified lipid species each. PGs, LacCer, SHexCer, PSs and HexCers, on the other hand, showed only few identified lipid species each (1, 2, 3, 4 and 4). Additionally, Cers, LPCs, PIs, LPEs, and FFAs were detected with 17, 16, 11, 8 and 8 identified lipid species, respectively. The dominance of identified lipid species within their respective lipid classes are shown in **Supplementary Fig. 1**. The coefficient of variation (%RSD) of peak areas was on average 17.41% for internal standards (**Supplementary Table 1**).

### Difference in the outcome between the diets

In total, 11 lipid species from phosphatidylcholine and sphingomyelin lipid classes and LPC(O-16:0) had different outcome between-groups (*p* < 0.05). In total, six out of the 83 annotated PCs, four out of the 30 annotated SMs, and LPC(O-16:0) had different outcomes. Lipid names, 95% CI, FDR p-values and the interaction slopes are given in **Table 2**. The strongest differences in the diet-outcomes were monounsaturated SM(d36:1): (β±SE: 1.44±0.28, *p* = 0.01) and polyunsaturated PC(P-36:4)/PC(O-36:5): (β±SE: 1.34±0.24, *p* = 0.01). All the aforementioned lipid species were present in significantly higher amounts after LCD than after HCD (see the “Slope-LCD” and “Slope-HCD” columns in **Table 2**).

**Table 2.** Results of the mixed-effects model. Shown in the table are (Name) of individual lipid species, interaction slope (Slope), its lower and upper confidence intervals (L95, U95), standard error, p-value of the slope (FDR) after correction for multiple testing, and slopes for LCD and HCD models (Slope-LCD, Slope-HCD).

| Name | Slope | L95 | U95 | SE | FDR | Slope-LCD | Slope-HCD |
| --- | --- | --- | --- | --- | --- | --- | --- |
| PC(P-36:4)/PC(O-36:5) | 1.34 | 0.83 | 1.85 | 0.24 | 0.009 | 0.63 | -0.70 |
| SM(d36:1) | 1.44 | 0.85 | 2.03 | 0.28 | 0.010 | 1.06 | -0.37 |
| PC(P-38:5)/PC(O-38:6) | 1.04 | 0.53 | 1.56 | 0.24 | 0.029 | 0.67 | -0.37 |
| SM(d36:2) | 0.76 | 0.38 | 1.13 | 0.18 | 0.029 | 0.65 | -0.10 |
| PC(P-36:2)/PC(O-36:3) | 1.59 | 0.77 | 2.42 | 0.39 | 0.036 | 0.89 | -0.70 |
| SM(d34:2) | 1.39 | 0.65 | 2.15 | 0.36 | 0.042 | 0.80 | -0.59 |
| PC(31:0) | 1.52 | 0.67 | 2.36 | 0.40 | 0.043 | 1.06 | -0.46 |
| PC(P-38:4)/PC(O-38:5) | 1.32 | 0.57 | 2.07 | 0.35 | 0.043 | 1.04 | -0.28 |
| PC(35:4) | 1.04 | 0.45 | 1.64 | 0.28 | 0.043 | 0.78 | -0.26 |
| LPC(O-16:0) | 1.28 | 0.55 | 2.01 | 0.35 | 0.043 | 0.32 | -0.95 |
| SM(d38:2) | 0.89 | 0.38 | 1.39 | 0.24 | 0.043 | 0.64 | -0.25 |

### Association between lipid species and clinical variables

Finally, we investigated how the 11 lipid species, which responded to the two diets in different ways, were associated with metabolic characteristics reported previously. In the linear mixed-effects model, there was an inverse association with the BMI and the lipid species PC(35:4): (β±SE: −0.24±0.05, *p* = 0.0005) and PC(31:0): (β±SE: −0.21±0.06, *p* = 0.003). The same trend of inverse association was also observed in diastolic BP, although not to statistical significance after correcting for multiple testing. PC(35:4) and PC(31:0) lipid species were elevated during the LCD intervention (see column “Slope-LCD” and “Slope-HCD” in **Table 2**), inferring a link between diet, circulating lipid concentrations, BMI and BP. There were no associations with the systolic BP that could be detected with statistical significance before and after correcting for multiple testing in this small study.

There was a positive association with the HDL-cholesterol and three poly-unsaturated phosphatidylcholine species (PC(35:4), PC(P-36:4)/PC(O-36:5), PC(P-38:4)/PC(O-38:5)) and two poly-unsaturated sphingomyelin species (SM(d34:2), SM(d36:2)). Lipid names, 95% CI, FDR p-values and slopes are given in **Supplementary Table 2**.

One of the 11 significantly changed lipid species was associated both with BMI, diastolic BP, and HDL, namely the poly-unsaturated phosphatidylcholine 35:4. **Fig. 1a** shows boxplot of semi-quantified pre-post concentration comparisons of PC(35:4) in each diet intervention. **Fig. 1b**, and **Fig. 1c** shows scatter plots of PC(35:4) associations with BMI, and HDL-cholesterol.

**Fig 1.**
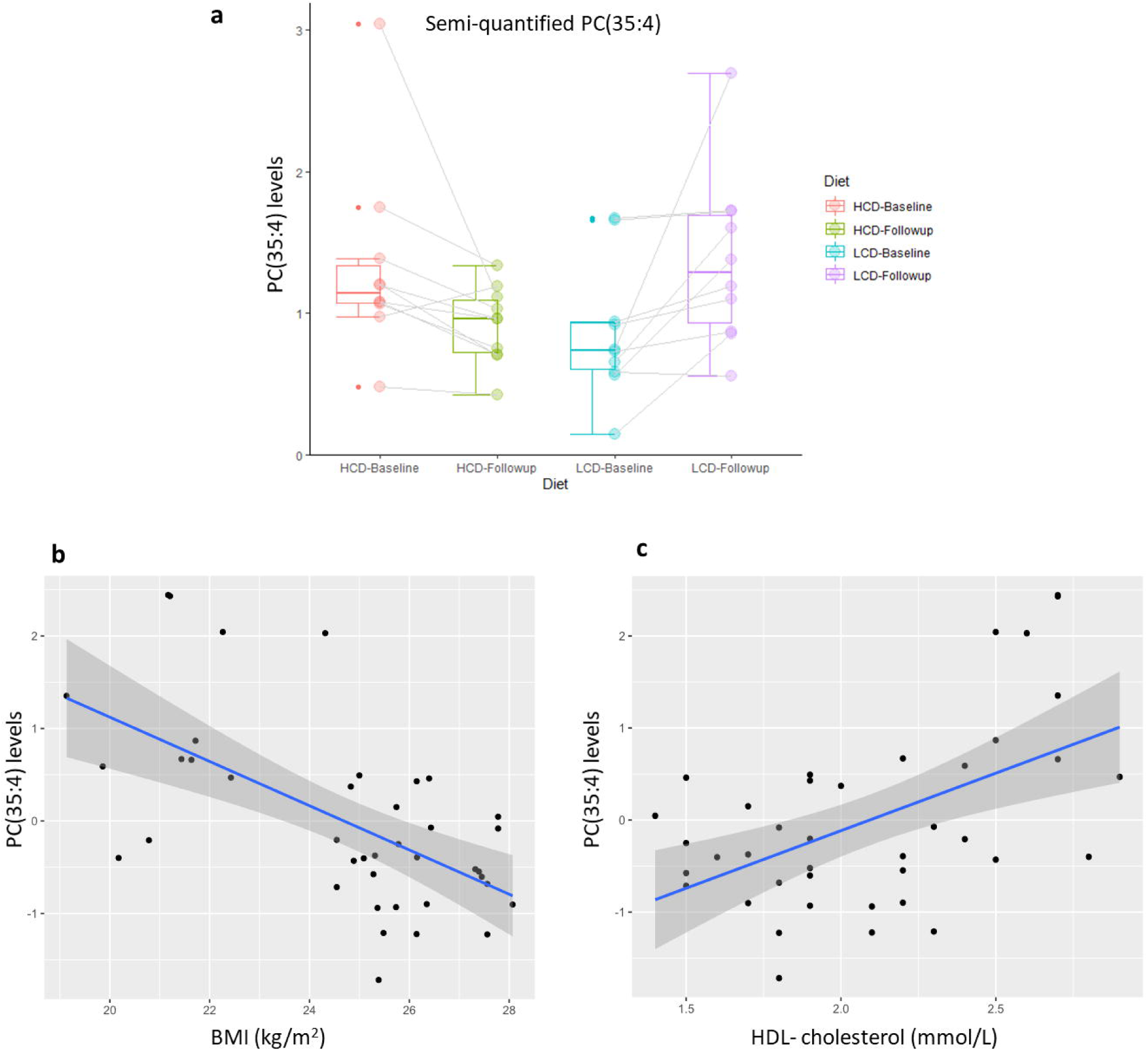
Jittered box plots of pre-post lipid levels comparisons of PC(35:4) in each diet intervention and its association with BMI and HDL-cholesterol. PC(35:4): (β±SE: 1.04±0.28, p= 0.043) was elevated during LCD (**a**) and inversely associated with BMI (**b**) and positively associated with HDL-cholesterol (**c**). **HB:** High carbohydrate diet at baseline, **HF:** High carbohydrate diet at follow-up, **LB:** Low carbohydrate diet at baseline, **LF:** Low carbohydrate diet at follow-up.

## Discussion

To our knowledge, this is the first study comparing the lipidome in individuals with type 1 diabetes before and after low carbohydrate diet and high carbohydrate diet. Lipids are biomolecules with important functions and their amount in the diet can modulate the blood lipidome in individuals with type 1 diabetes. In this study, we performed lipidomics in a randomized cross-over trial and we compared the fasting plasma lipid profiles from individuals with type 1 diabetes before and after ingesting isocaloric LCD and HCD.

The main result of this study was that when both LDC and HDC were compared, 11 lipid species belonging to sphingomyelin and phosphatidylcholine were significantly changed in outcome between the diets (**Table 2**). Additionally, and most importantly, when we studied each trial arm separately, we observed that all the 11 significantly changed lipid species in the outcome were in greater concentration in the LCD arm, but none were altered significantly in the HCD arm. This was an unexpected result since total cholesterol, LDL and triglycerides had not changed significantly in this trial and only HDL-cholesterol was significantly elevated in the LCD arm, suggesting a moderate lipid improvement with LCD. Also this suggests that a prolonged LCD could help with adjusting lipid classes since there is existing evidence that individuals with type 1 diabetes have decreased levels of blood sphingolipids and phosphatidylcholines, abundant in HDL [26].

Phosphatidylcholines and sphingomyelins are two important classes of phospholipids essential for cell membrane function and major components of HDL-cholesterol specific those containing polyunsaturated fatty acids [14, 27,28]. Interestingly a noteworthy result from our study is that the LCD arm showed that HDL-cholesterol was increased and positively associated with polyunsaturated sphingomyelins and phosphatidylcholines (**Supplementary Table 2**). In rodents, sphingomyelin supplementation has been shown to help reduce the intestinal absorption of serum lipids such as cholesterol and triglycerides [12]. Sphingomyelin levels have also been associated with mild-to-moderate hypertension [29]. Based on these findings, supplementation could be an avenue to explore in the context of dyslipidemia in type 1 diabetes. The findings for this study were in line with previous work on LCD in which similarly HDL-cholesterol and diastolic BP were increased and decreased respectively with less carbohydrate intake, in a cross-sectional carbohydrate intake study (carbohydrate intake of <130 g/day and >253 g/day) in type 1 diabetes [2]. Analogous results have been reported in individuals with type 2 diabetes, in which LCD resulted in a significant increase in HDL-cholesterol and decrease in high BP [30,31].

Previous studies have demonstrated that abnormal sphingolipids levels are found at the onset of type 1 diabetes in regulating beta cell biology and inflammation [26]. Fenofibrate, a lipid lowering medication is known to regulate sphingolipid metabolism and has been suggested as an important treatment in the management of dyslipidaemia [32]. It has been shown that very-long-chain sphingomyelin were increased in three weeks fenofibrate-treated NOD mice and this had a beneficial effect on blood glucose homeostasis [32]. Interestingly, the main study outcome from Schmidt et al. [18] showed that glycemic variability and time spent in hypoglycemia were lower during LCD than during HCD.

As the total amounts of cholesterol did not change with LCD, this was a positive result since hypercholesterolemia is a major risk factor for developing cardiovascular disease [33-35]. The presence of CVD is related to disturbances in lipoprotein metabolism and dyslipidemia [36]. Dyslipidemia in individuals with diabetes not only fuels the reduction of HDL-cholesterol concentration, but also it modifies the composition of lipoprotein [3-6]. Increasing evidence suggests that in patients with chronic inflammatory disorders, HDL-cholesterol may lose important antiatherosclerosis properties and become dysfunctional [36]. So far, no therapeutic strategy to raise HDL-cholesterol has been successful in reducing CVD [36]. In this study we found three phosphatidylcholine lipid species (PC-(O-36:5), PC-(O-38:5) and PC(35:4)) and two sphingomyelin lipid species (SM(d36:2) and SM(d34:2)) associated with HDL-cholesterol (**Supplementary Table 2**). Interestingly all these lipid species were increased during the LCD-arm suggesting that LCD led to better functionality and composition of HDL-cholesterol particles as they are enriched in plasma phosphatidylcholine (**see the “Slope-LCD” column in Table 2**).

Remarkably, a previous lipidomics study in 3779 type 2 diabetes-based cohort researchers found that PC(35:4) lipid species out of the 7 novel lipid species were associated with CVD [37]. PC(35:4) was suggested to improve prediction of CVD mortality. Remarkable result to emerge from our data is that, PC(35:4) was the strongest positively associated lipid species with HDL-cholesterol (**Supplementary Table 2**) and increased during the LCD-arm. Importantly, PC(35:4) was also inversely associated with BMI (**Fig. 1**.**b**) and Diastolic BP. This and our investigation demonstrate the potential of lipid species as biomarkers for CVD risk in type 1 diabetes.

### Strengths and limitations

There are certain limitations in the present study: Exact food intake for each study participant was not recorded. The diet plans were not conducted in a controlled environment. This means that it is difficult to know, what proportion of other energy sources ╌ proteins or fats╌ the carbohydrates were replaced with in the LCD. While this varies by participant, we now know that the replacement resulted in the elevation of the concentrations of sphingomyelins and phosphatidylcholines in LCD. Another limitation is the small sample size of 10 participants, which, was mitigated by the crossover study design. We view the randomized crossover study design and the high level of adherence to the carbohydrates criteria during LCD and HCD as two core strengths of this study. Further, the comprehensive lipidomic profiling is a strength of this study in type 1 diabetes.

In conclusion, our novel data now provide the foundation for future work aimed at diet lipid modulation in type 1 diabetes. We have demonstrated that low carbohydrate diet elevates blood sphingomyelin and phosphatidylcholine lipid species in type 1 diabetes, which are thought to reduce dyslipidaemia. Further, one single lipid species, poly-unsaturated phosphatidylcholine 35:4 was inversely associated with BMI and positively associated with HDL-cholesterol. Therapeutic development for diabetic complications such as dyslipidemia and cardiovascular disease will require a better understanding of the crosstalk between diet and lipid metabolism in individuals with type 1 diabetes. This study provides support for several existing paradigms of dyslipidaemia and suggests new avenues of prevention of dyslipidaemia in type 1 diabetes. Confirming our findings, dietary lipid species, such as PC(35:4) should be tested in a larger cohort to fully understand the therapeutic opportunities in type 1 diabetes.

## Data Availability

personal sensitive data can not be deposited

## Acknowledgements

We acknowledge BioRender for providing the online tool which allowed us to design the graphical abstract for our paper.

## Data availability

All the lipidomics data generated and analyzed in this study are included in this manuscript and its supplementary files.

## Funding

This study did not receive any funding, but the original trial was funded by a research grant from the Danish Diabetes Academy supported by the Novo Nordisk Foundation, the Danish Diabetes Association, Vissing Fonden, and The A.P. Møller Foundation.

## Duality of interest

None of the investigators has personal interests in the conduct or the outcomes of the study.

## Contribution statement

C.L-Q., K.N. and F.P. designed and supervised the study. S.S., A.R. and M.B.C. undertook the clinical study. N.A. performed lipidomics analysis, statistical analysis and wrote the manuscript. T.S. and M.K. planned and supervised the statistical analysis. All authors critically reviewed and approved the final manuscript.

## Abbreviations

Cers: Ceramide species
FFAs: Free fatty acid species
HexCer: Hexosyl-ceramide species
LacCer: Lactosyl-ceramide
LCD: Low carbohydrate diet
LPCs: Lyso-phosphatidylcholine species
LPC-Os: Lyso-alkyl-phosphatidylcholine species
LPEs: Lyso-phosphatidylethanolamines
HCD: High carbohydrate diet
NOD: Non-obese diabetic
NIST: National Institute of Standards and Technology
PCs: Phosphatidylcholine species
PC-Os: Alkyl-acyl-phosphatidylcholine species
PGs: Phosphatidylglycerol species
PEs: Phosphatidylethanolamine species
PIs: Phosphatidylinositol species
PSs: Phosphatidylserine species
SHexCer: Sulfatide species
SMs: Sphingomyelin species
TGs: Triacylglycerol species

## Notes

### Competing Interest Statement

The authors have declared no competing interest.

### Clinical Trial

NCT02888691

### Funding Statement

The lipidomics study did not receive any particular funding. The clical trial was funded by a research grant from the Danish Diabetes Academy supported by the Novo Nordisk Foundation, the Danish Diabetes Association, Vissing Fonden, and The A.P. Moller Foundation.

### Author Declarations

The study protocol was approved by the Regional Scientific Ethics Committee (H-16032632), the Danish Data Protection Agency (AHH-2015-002), and registered at ClinicalTrials.gov (NCT02888691).

